# Deep Learning-based Real-Time Seizure Detection and Multi-Seizure Classification on Pediatric EEG

**DOI:** 10.1101/2024.11.25.24317933

**Authors:** Hyewon Jeong, Kwanhyung Lee, Seyun Kim, Hoon-Chul Kang, Donghwa Yang

## Abstract

**Background and Objective:** To develop a reliable and accurate seizure detection method using deep learning models capable of detecting and classifying multiple seizure types in real time.

**Methods:** We retrospectively collected electroencephalography (EEG) recordings, which were acquired as part of routine diagnostic tests for patients aged 3 months to ≤18 years of age with childhood absence epilepsy, infantile epileptic spasms syndrome, other generalized epilepsy, and focal epilepsy, between January 2018 and December 2022 at Severance Children’s Hospital. We used EEG recordings from both seizure and non-seizure patients, which were downsampled to 200 Hz for real-time seizure detection and multi-classification.

**Results:** Of the 199 patients (620 seizures), 49 (297 seizures) belonged to the childhood absence epilepsy group, 16 (200 seizures) to the infantile epileptic spasms syndrome group, 14 (76 seizures) to other generalized epilepsy group, 19 (47 seizures) to focal epilepsy group, and 101 to the normal group. The results showed the best overall performance of AUROC 0.98 and APROC of 0.73 with ResNet with Long-Short Term Network and a 12 s sliding window on real-time seizure detection task. Furthermore, ResNet50 without the frequency bands feature extractor showed the best overall weighted performance for multi-class seizure detection with 0.99 AUROC and 0.99 APPRC.

**Discussion:** Our approach proposes robust methods which include EEG preprocessing strategy with real-time detection/classification of multiple seizures, which helps monitor pediatric seizure. The result shows that real-time seizure detection can be effectively applied to real-world clinical datasets from a pediatric epilepsy unit with realistic performance and speed.

## INTRODUCTION

Since the advent of several powerful modern machine learning and deep learning models, several attempts have been made to detect seizures and abnormal waves in electroencephalography (EEG) signals.^1–5^ More recent studies have demonstrated the performance of advanced tasks such as multi-class seizure detection,^6–10^ and real-time seizure detection.^11–15^ Although previous studies have explored multiple model architectures for seizure detection and classification, certain limitations make their direct use in neurology wards challenging. First, very few studies were designed for real-time seizure detection,^11,12^ which limits their practical application. Moreover, several recent studies have focused on classifying pre-cut seizure signals, which is an approach with limited applicability in real-world clinical settings for seizure detection.^16–18^ Real-time seizure detection and classification are invaluable as timely identification allows for immediate intervention, which potentially aids in mitigating the severity of the episode. Furthermore, it also helps in preventing further complications and swift adjustment of treatment strategies.^19^

The performances of several previous seizure detection models^1–5^ have only been evaluated using public benchmark datasets.^12, 20–23^ Furthermore, the reported model performances could not be fairly compared with each other as they used distinct experimental settings and evaluation metrics until Lee et al.^11^ proposed a comprehensive analysis of seizure detection in real-time setting.

Developing and deploying real-time seizure detectors and classifiers is crucial for the diagnosis and treatment of patients with epilepsy. Seizure events of patients are routinely monitored with 24h inpatient video-EEG recordings. Onset of seizure is usually recorded by pressing the button when the family members notice the onset of ictal period that could be of noticeable clinical importance. However, several event onsets are prone to be missed on-site. This might be due to the fact that medical associates cannot always monitor the EEG signals. Moreover, even family members cannot always notice the onset of seizures for some types of seizures which have subtle clinical symptoms, such as infantile epileptic spasms syndrome.

This study aimed to investigate the applicability of our deep learning-based real-time seizure detection model to real-world pediatric EEG datasets and validate its generalizability for multiple seizure classifications. We used the models from the study by Lee et al.,^11^ that showed promising results for real-time seizure detection tasks when applied to public adult EEG data with seizure events.

In this study, the models constructed for real-time seizure detection incorporated the architectures of Convolutional Neural Networks (CNN) and autoregressive layers. The configurations included were CNN2D+Long-Short Term Network (LSTM), ResNet-short+LSTM, ResNet-short+Dilation+LSTM, and MobileNetV3-short+LSTM. Moreover, for the multi-class classification task, we explored multiple CNN baselines such as ResNet50, MobilenetV3, and DenseNet. We extended the binary real-time seizure detection of Lee et al.^11^ to multi-seizure classification tasks, and evaluated the performance of the two tasks on pediatric EEGs within a cohort of four types of seizures: Childhood Absence Epilepsy (CAE), Infantile Epileptic Spasms Syndrome (IESS), Generalized Epilepsy (GN), and Focal Epilepsy (FC). The study findings would exemplify the applicability and generalizability of our approach for readily applicable deep learning-based real-time seizure detection and multi-seizure classification.

## METHODS

### Study population

This study was performed at the epilepsy clinic of Severance Children’s Hospital between 2018 and 2022, and included patients under the age of 18 years with seizures. The inclusion criteria were: (1) patients diagnosed with CAE, IESS, other generalized epilepsy, and focal epilepsy; and (2) those who had seizures during EEG. We excluded patients who did not have seizures during EEG recording. In addition, we obtained EEG records from patients with headache who did not have any seizure episodes, and designated them as the negative control group.

All the procedures in this study were performed in accordance with the tenets of Declaration of Helsinki. The study was approved by the Institutional Review Board of Severance Hospital (IRB 4-2023-0967).

### Data collection

We reviewed the patients’ medical records and collected data regarding age of seizure onset, age at the time of diagnosis, brain magnetic resonance imaging (MRI) findings, number of anti-seizure medications, EEG findings, and raw EEG data. First, we analyzed EEG data from patients diagnosed with CAE, IESS, other generalized epilepsy, and focal epilepsy, ensuring that their seizures were captured during the EEG recordings. Following this, two pediatric neurologists annotated the data, marking the onset times of seizures for each patient.

### Dataset Preprocessing

We developed a Python program that utilized the Minimum-norm estimation (MNE) open-source package to facilitate preprocessing and extraction of EEG data from files with edf extensions. To implement real-time seizure detection, our first step involved identifying specific seizure periods [Figure 1(a), Step 1]. Subsequently, we bracketed each seizure period with up to 1h of non-seizure period on either side of the identified seizure. These data were then segmented into non-overlapping multiple 30 s intervals of EEG signals, which formed the basis for our training set. We excluded EEG signal data chunks with lengths of less than 30 s. To streamline the training process for real-time detection, we randomly chose a maximum of ten 30 s non-seizure intervals per one 30 s seizure interval.

**Figure 1.**
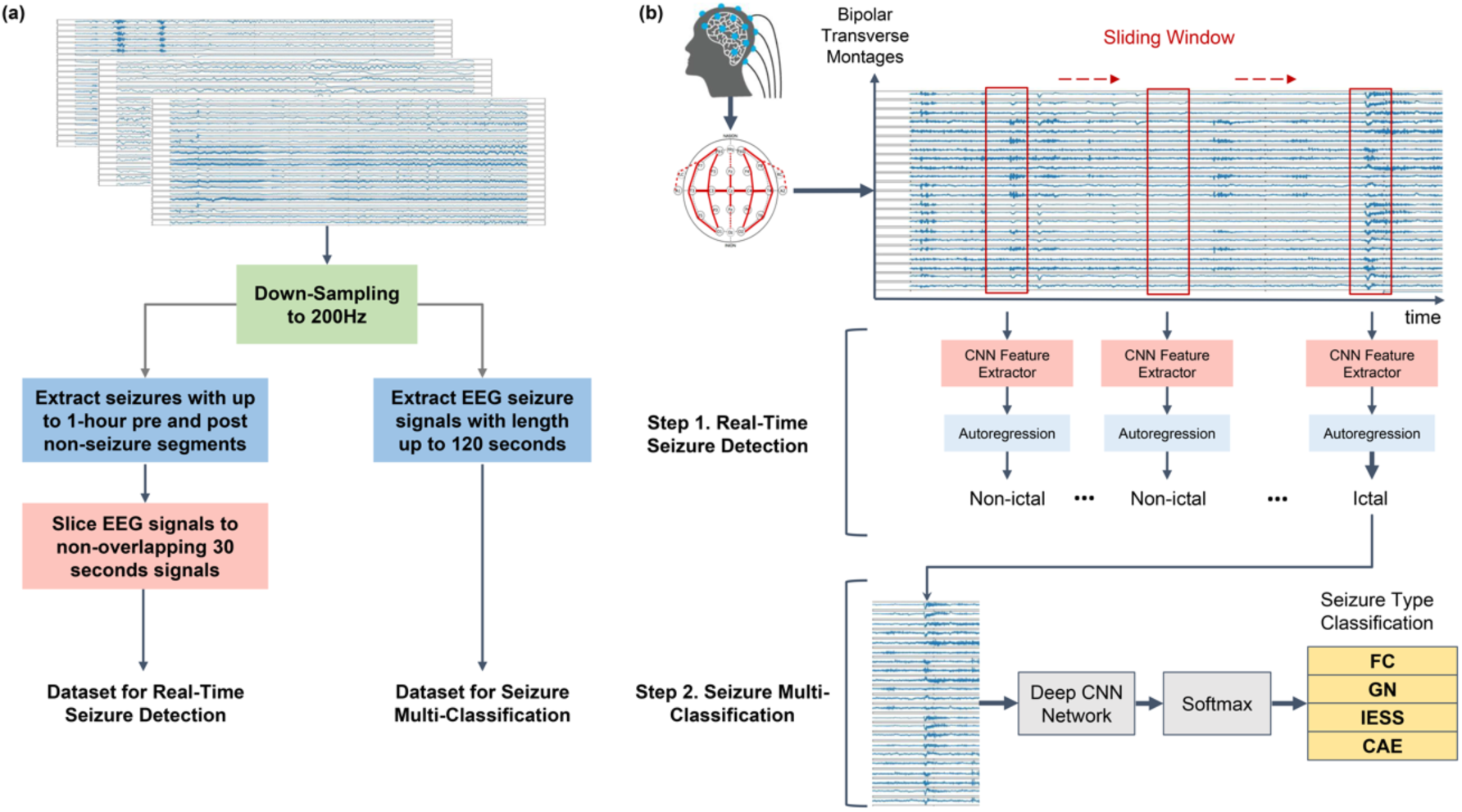
Schematic illustration of (a) seizure signal preprocessing and real-time seizure detection [(b) Step 1], multi-class seizure classification process [(b) Step 2]. (a) Preprocessing for seizure detection and classification. We segmented the entire seizure signal into shorter 30 s segments, creating a chunked dataset. Additionally, for multi-class classification, the dataset included up to 30 s of non-ictal period prior to the seizure onset (total up to 120 s), enriching the model with valuable pre-ictal signal information. (b) Two-stage seizure detection and classification. We identify the seizure using the sliding window (Step 1. real-time seizure detection) and classified the detected seizures into one of the multiple possible types (Step 2. multi-class classification).

**Figure 2.**
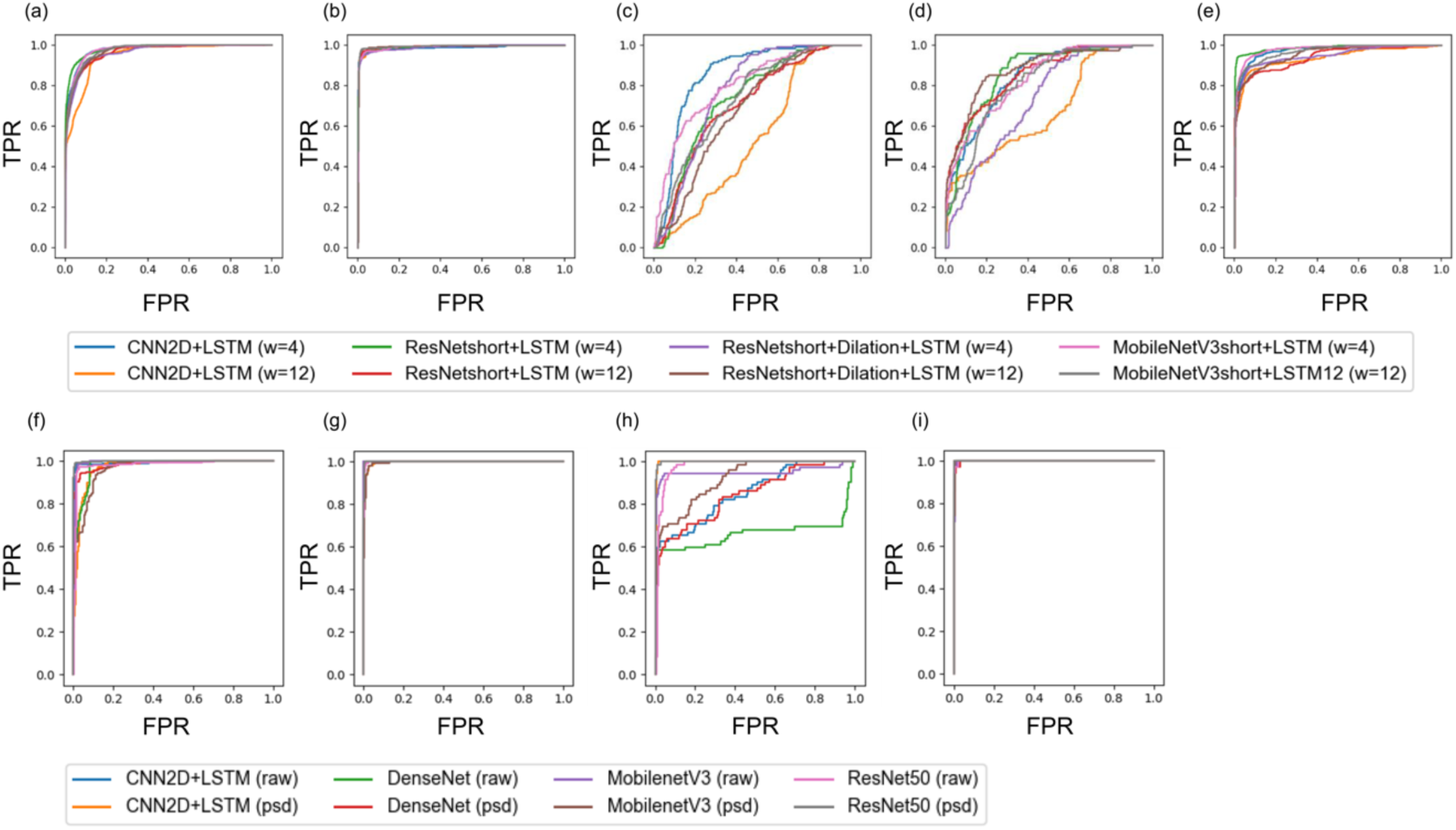
Subgroup Area Under the Curve (AUROC) plots for binary seizure [Figures (a)-(e)] and multi-class seizure detection [Figures (f)-(i)]. Figures (a)-(e) represent the AUC for detecting various seizure types: (a) any type, (b) CAE, (c) IESS, (d) GN, and (e) FC. Figures (f)-(i) illustrate the AUC for detecting different seizure types in multi-class classification tasks: (f) CAE, (g) IESS, (h) GN, and (i) FC. W=4 and W=12 indicates window length of 4 and 12 s, respectively. For multi-class seizure classification model, we included the cases of using raw signal (raw) and power spectral density (frequency bands, psd) were used.

For multi-seizure classification, we identified the specific time frames during which the seizures occurred. To gather information regarding transition from non-seizure to seizure periods, we encapsulated the identified seizure segments with non-seizure durations. These comprised non-ictal periods of a maximum of 30 s before and up to 5 s after each identified maximum 120 s seizure episode. In cases where the non-ictal period preceding the targeted seizure period was shorter than 30 s due to a previous seizure or absence of a previous recording, the maximum possible non-ictal period was collected for multi-class seizure classification input. For EEG signals extending beyond 120 s, we confined our analysis to the initial 120 s. Separately, for multi-class seizure classification task, we defined the input seizure EEG signal to be ≤ 120 s, as we were more concerned about classifying seizure types than fast real-time detection.

For both real-time seizure detection and multi-seizure classification, we sampled raw EEG recordings at a sampling rate of 200 Hz, the lowest frequency recorded in our pediatric patient population. We used double banana montage that included transverse montage added to bipolar montage. This montage included the following electrode pairs: ‘fp1-f7’, ‘fp2-f8’, ‘f7-t3’, ‘f8-t4’, ‘t3-t5’, ‘t4-t6’, ‘t5-o1’, ‘t6-o2’, ‘t3-c3’, ‘c4-t4’, ‘c3-cz’, ‘cz-c4’, ‘fp1-f3’, ‘fp2-f4’, ‘f3-c3’, ‘f4-c4’, ‘c3-p3’, ‘c4-p4’, ‘p3-o1’, ‘p4-o2’, ‘fz-cz’, and ‘cz-pz’. Differences between the signals from two adjacent leads were extracted to obtain these electrode pairs. The amplitude of each seizure was normalized on the basis of minimum and maximum values of training data signals.

For our main result, we did not further use signal feature extractors. This was due to the fact that Lee et al.^11^ demonstrated that the seizure detection model that used raw dataset without any signal feature extractors performed at par with those with signal feature extractors. We also tested the Frequency Bands feature extractor introduced by Lee et al.,^11^ specifically for multi-seizure classification tasks. The extractor averaged the signal data within the frequency ranges of 1–4, 4–8, 8–12, 12–30, 30–50, 50–70, and 70–100 Hz. According to Lee et al.,^11^ this extractor achieved the second fastest processing speed and showed a relatively high seizure detection performance among the five feature extractors. For real-time seizure detection, the model obtained an input of 30 s signal data at each epoch, wherein a shorter sliding window shift. We explored the performance of four different models with 4 s^11^ and 12 s^11, 24^ sliding window lengths and 1 s window shift length. For multi-class classification task, the model obtained the entire cropped signal chunk with the label for whole signal, indicating whether an input chunk included each corresponding seizure type.

Each input window was labeled with a categorical seizure label for multi-class seizure classification. Each window was labeled with five categorical labels according to the International League Against Epilepsy (ILAE) classification:^25^ normal signal, CAE, IESS, GN, and FC. The data processing steps explained in this section followed the same settings as those in our previous study.^11^

### Model Specification

We extensively compared the performance of the models for real-time seizure detection and multi-class seizure classification tasks. For real-time detection networks, we adopted the model code and training loop of real-time seizure detection from a previous study^11^ (https://github.com/AITRICS/EEG_real_time_seizure_detection). Among the 15 models tested by Lee et al.,^11^ we selected four lightweight and fast models (CNN2D+LSTM, ResNet-short+LSTM, ResNet-short+Dilation+LSTM, and MobileNetV3-short+LSTM) that were at par with the best performing model. All models implemented for real-time seizure detection integrated the architecture of CNN with autoregressive models, as illustrated in Figure 1 (b).

For multi-class seizure classification, the following five models were tested: CNN2D+LSTM,^11^ ResNet50+Conv2D, DenseNetV3,^26^ and MobileNetV3.^27^ We adapted these models from their original designs with minor alterations to the CNN modules, specifically employing a one-dimensional filter for improved signal information extraction. The training process utilized a batch size of 16 and was trained for 20 epochs using the Adam optimizer with a learning rate sweep ranging between 1e^(−3) - 1e^(−5). The model was optimized using binary cross-entropy loss function for real-time seizure detection, and cross-entropy loss for multi-classification task.

### Model Evaluation

To evaluate the model’s performance, we conducted 5-fold cross-validation and calculated the average Area Under the Receiver Operator Curve (AUROC) and Area under the Precision Recall Curve (AUPRC) scores. We evaluated the performance of each experimental setting and model by comparing their performances. The models were evaluated using the scikit-learn module.^28^

## RESULTS

### Baseline characteristics

A total of 98 patients were diagnosed with epilepsy and treated with anti-seizure medications between 2018-2022. One hundred and one patients with headache had normal EEG findings and were classified as the negative control group. Finally, 199 patients were included in this study.

The median age of the patients at the time of diagnosis was 7.7 years (interquartile range, [IQR] 5.2-9.7), and the study population comprised 89 males (44.7%). Twenty-eight (51.9%) of the 54 patients had normal brain MRI findings. The IESS group was the youngest at the time of onset of seizures, and the proportion of female patients with CAE was high. There were no significant differences among the four groups in terms of baseline characteristics, including the number of anti-seizure medications (Table 1-1).

**Table 1-1.** Patient demographics stratified by epilepsy type.

| Variables | Total (n=199) | Normal (n=101) | CAE (n=49) | IESS (n=16) | Generalized (n=14)<sup>a</sup> | Focal (n=19) |
| --- | --- | --- | --- | --- | --- | --- |
| Sex, male | 89 (44.7) | 50 (49.5) | 13 (26.5) | 10 (62.5) | 8 (57.1) | 8 (42.1) |
| Age at diagnosis, year | 7.7 (5.2-9.7) | 8.8 (12.3-14.7) | 7.5 (5.5-8.6) | 0.6 (0.5-0.9) | 10.6 (7.9-13.0) | 12.4 (9.4-15.8) |
| Age at seizure onset, year | 5.5 (1.9-7.6) | - | 6.2 (4.9-7.6) | 0.6 (0.4-0.8) | 2.2 (1.1-7.4) | 8.1 (4.0-9.4) |
| Normal brain MRI findings | 28 (51.9) [54]<sup>b</sup> | - | 5 (100.0) [5] | 8 (50.0) | 8 (57.1) | 7 (36.8) |
| Number of ASM | 2 (2-4) | - | 2 (1-2) | 2 (2-4) | 4 (4-5) | 4 (3-4) |

values are presented as median (interquartile range) or n (%). CAE, childhood absence epilepsy; IESS, infantile epileptic spasms syndrome; ASM, antiseizure medication.

### EEG Dataset for Seizure Detection and Multi-class Seizure Classification

We have summarized the number of patients, length of the negative signal (negatively labeled for seizures, i.e., a signal without seizure events), and positive signal (signal labeled positive to contain seizure events) for each seizure type in Table 1-2. We included a total of 199 patients, including 101 healthy individuals, 98 patients with childhood epileptic seizure (620 seizures), 49 with CAE (297 seizures), 16 with IESS (200 seizures), 14 with GN (76 seizures), and 19 with FC (47 seizures) (Table 1-2).

**Table 1-2.** Characteristics of EEG data utilized for binary real-time detection and multi-class seizure classification tasks.

| Group | Dataset Type | Number of Patients | Negative Length (sec) | Positive Length (sec) | Average Event Length (sec) | Number of Seizure |
| --- | --- | --- | --- | --- | --- | --- |
| Normal | Train | 81 | 136,110 | 0 | N/A | N/A |
|  | Test | 20 | 34,830 | 0 | N/A | N/A |
| CAE | Train | 40 | 55,377 | 2,402 | 9.2 | 262 |
|  | Test | 9 | 12134 | 376 | 10.74 | 35 |
| IESS | Train | 13 | 88304 | 157 | 0.95 | 166 |
|  | Test | 3 | 17600 | 33 | 0.98 | 34 |
| Generalized | Train | 12 | 29313 | 532 | 7.95 | 67 |
|  | Test | 2 | 3552 | 77 | 8.61 | 9 |
| Focal | Train | 16 | 51506 | 3358 | 83.96 | 40 |
|  | Test | 3 | 10397 | 814 | 116.33 | 7 |

<sup>a</sup> “Generalized” is classified except the absence of seizure and epileptic spasms
<sup>b</sup> n in brackets means the number of patients examined.

### Real-Time Seizure Detection

For real-time binary seizure detection, we first chose a short (4 s) and a relatively long sliding window length (12 s) for input of four distinct models, and analyzed the performance of each model (Table 2). A window size of 4 s was the optimal setting chosen by Lee et al. (2022), considering the trade-off between performance and speed. The 12 s sliding window was the closest window length to the size of the physicians process each time (15 s). The ResNet-short + LSTM model with a 12 s sliding window demonstrated the best overall performance for real-time seizure detection among the four different models with two different window lengths, demonstrating an AUC of 0.9799 for detecting any kind of seizure (Table 2).

**Table 2.** Performance of binary real-time seizure detection with 4 different models and window sizes of 4 and 12 s. We summarize the 5-fold cross-validation test results, with the average performance and corresponding standard deviations (std) for each model and window size.

| Window Size (Seconds) |  | 4 |  |  | 12 |  |  |
| --- | --- | --- | --- | --- | --- | --- | --- |
| Models | Seizure type | AUC (std) | APR (std) | F1 (std) | AUC (std) | APR (std) | F1 (std) |
| CNN2D + LSTM | Any | 0.9727<br>(0.00) | 0.7027<br>(0.04) | 0.6399<br>(0.05) | 0.9687<br>(0.00) | 0.7031<br>(0.04) | 0.6641<br>(0.06) |
|  | CAE | 0.9888<br>(0.00) | 0.9743<br>(0.01) | 0.9092<br>(0.02) | 0.9952<br>(0.00) | 0.9873<br>(0.00) | 0.9445<br>(0.00) |
|  | IESS | 0.8721<br>(0.01) | 0.5552<br>(0.02) | 0.2246<br>(0.05) | 0.7988<br>(0.05) | 0.6317<br>(0.05) | 0.3708<br>(0.09) |
|  | Generalized | 0.8347<br>(0.05) | 0.648<br>(0.04) | 0.313<br>(0.09) | 0.8117<br>(0.04) | 0.6371<br>(0.03) | 0.3092<br>(0.04) |
|  | Focal | 0.953<br>(0.02) | 0.9029<br>(0.03) | 0.7432<br>(0.03) | 0.9394<br>(0.02) | 0.8933<br>(0.04) | 0.7432<br>(0.07) |
| ResNet-short + LSTM | Any | 0.9684<br>(0.00) | 0.6884<br>(0.04) | 0.6468<br>(0.02) | 0.9799<br>(0.00) | 0.7302<br>(0.03) | 0.6746<br>(0.02) |
|  | CAE | 0.9877<br>(0.00) | 0.9684<br>(0.01) | 0.9113<br>(0.01) | 0.9945<br>(0.00) | 0.9826<br>(0.01) | 0.9345<br>(0.02) |
|  | IESS | 0.7955<br>(0.04) | 0.5282<br>(0.01) | 0.1368<br>(0.02) | 0.7454<br>(0.04) | 0.5594<br>(0.03) | 0.2461<br>(0.05) |
|  | Generalized | 0.757<br>(0.09) | 0.6103<br>(0.03) | 0.2643<br>(0.05) | 0.8578<br>(0.03) | 0.7151<br>(0.04) | 0.4661<br>(0.07) |
|  | Focal | 0.957<br>(0.01) | 0.9122<br>(0.01) | 0.782<br>(0.02) | 0.9463<br>(0.02) | 0.8954<br>(0.05) | 0.746<br>(0.1) |
| ResNet-short+Dilation + LSTM | Any | 0.9679<br>(0.01) | 0.6855<br>(0.02) | 0.6497<br>(0.03) | 0.9678<br>(0.01) | 0.7227<br>(0.04) | 0.6643<br>(0.04) |
|  | CAE | 0.9891<br>(0.00) | 0.9578<br>(0.03) | 0.887<br>(0.05) | 0.994<br>(0.00) | 0.9766<br>(0.02) | 0.9083<br>(0.08) |
|  | IESS | 0.8229<br>(0.03) | 0.5344<br>(0.01) | 0.6813<br>(0.03) | 0.7285<br>(0.03) | 0.5487<br>(0.02) | 0.222<br>(0.04) |
|  | Generalized | 0.7575<br>(0.05) | 0.6192<br>(0.03) | 0.3219<br>(0.05) | 0.836<br>(0.01) | 0.6607<br>(0.04) | 0.3616<br>(0.05) |
|  | Focal | 0.9544<br>(0.5) | 0.9084<br>(0.03) | 0.775<br>(0.05) | 0.9576<br>(0.01) | 0.926<br>(0.03) | 0.806<br>(0.07) |
| MobileNetV3-short + LSTM | Any | 0.9675<br>(0.00) | 0.6921<br>(0.05) | 0.6415<br>(0.05) | 0.9627<br>(0.02) | 0.6724<br>(0.07) | 0.6264<br>(0.06) |
|  | CAE | 0.9881<br>(0.00) | 0.9724<br>(0.00) | 0.8992<br>(0.01) | 0.9932<br>(0.00) | 0.9785<br>(0.01) | 0.9155<br>(0.02) |
|  | IESS | 0.7597<br>(0.06) | 0.5243<br>(0.02) | 0.113<br>(0.03) | 0.6411<br>(0.12) | 0.5404<br>(0.04) | 0.18<br>(0.06) |
|  | Generalized | 0.7735<br>(0.04) | 0.6183<br>(0.02) | 0.2975<br>(0.04) | 0.8094<br>(0.09) | 0.6626<br>(0.08) | 0.36<br>(0.13) |
|  | Focal | 0.9684<br>(0.03) | 0.9272<br>(0.02) | 0.7902<br>(0.05) | 0.9594<br>(0.09) | 0.9136<br>(0.08) | 0.8092<br>(0.06) |

The performances of the four real-time seizure detection models demonstrated gaps across the distinct seizure-type subgroups. Real-time detection of CAE showed superior performance over detection of any type (Any in Table 2, AUC of 0.9799 with ResNet-short + LSTM, 12 s window), achieving an AUC of 0.9945 with ResNet-short + LSTM using a window size of 12 s (CAE in Table 2). Detecting focal seizures (Focal in Table 2) showed a slightly lower performance compared to detecting any kind of seizure (Any in Table 2), with an AUC of 0.957 with the same window size (12 s) and model architecture (ResNet-short + LSTM). Finally, the detection of IESS and generalized seizures exhibited the lowest performance among the four seizure types, with AUCs of 0.7454 and 0.8578, respectively, when using the same window size (12 s) and model architecture (ResNet-short + LSTM).

Among the four models that were evaluated, increasing the window size led to an improved performance for CAE detection (Table 2, ResNet-short + LSTM model showed an AUC of 0.9877 for a window size of 4 s and 0.9945 for 12 s), while resulting in decreased performance for IESS detection (ResNet-short + LSTM model showed an AUC of 0.7955 for 4 s and 0.7454 for 12 s). The performance of detecting focal seizures was not significantly affected by the size of the sliding window (ResNet-short + LSTM model showed an AUC of 0.957 for 4 s and 0.9463 for 12 s window length). The real-time seizure detection models employing deep hidden layers with a 12 s sliding window (ResNet-short + LSTM, ResNet-short + Dilation + LSTM, and MobileNetV3-short + LSTM) exhibited higher seizure detection performance in comparison to a relatively shallow hidden layer model (CNN2D + LSTM).

### Multi-Class Seizure Classification

We have presented the performances of four different models with and without frequency bands feature extractor for multi-class seizure detection in Table 3. Among the four models, ResNet50 without the frequency bands feature extractor exhibited the best overall performance, with a weighted AUC of 0.9961, APR of 0.9912, and F1 score of 0.9689.

**Table 3.** Performance of multi-class seizure classification model. We summarize the 5-fold cross validation test results with the averaged performance and corresponding standard deviations (std).

| Extractor Type |  | Raw |  |  | Frequency Bands |  |  |
| --- | --- | --- | --- | --- | --- | --- | --- |
| Mod<br>els | Type | AUC (std) | APR (std) | F1 (std) | AUC (std) | APR (std) | F1 (std) |
| CN<br>N2D<br>+<br>LST<br>M | CAE | 0.9976<br>(0.0032) | 0.996<br>(0.0056) | 0.9805<br>(0.0204) | 0.8865<br>(0.0627) | 0.8859<br>(0.0609) | 0.8375<br>(0.0475) |
|  | IESS | 1.0 (0.0) | 1.0 (0.0) | 0.9886<br>(0.0165) | 0.9991<br>(0.0018) | 0.9986<br>(0.0028) | 0.9649<br>(0.0433) |
|  | Generalize<br>d | 0.8807<br>(0.1231) | 0.7258<br>(0.1372) | 0.7844<br>(0.1401) | 0.9789<br>(0.0257) | 0.8799<br>(0.0937) | 0.8187<br>(0.1544) |
|  | Focal | 0.9517<br>(0.034) | 0.7954<br>(0.0847) | 0.77<br>(0.0818) | 0.9571<br>(0.0422) | 0.6788<br>(0.2759) | 0.7028<br>(0.1542) |
|  | Weighted | 0.9824<br>(0.0124) | 0.9525<br>(0.0145) | 0.9457<br>(0.0269) | 0.9471<br>(0.0295) | 0.9132<br>(0.0497) | 0.8754<br>(0.0429) |
| Res<br>Net5<br>0 | CAE | 0.9944<br>(0.007) | 0.9937<br>(0.0071) | 0.9827<br>(0.0141) | 0.98<br>(0.032) | 0.9429<br>(0.049) | 0.9574<br>(0.0083) |
|  | IESS | 1.0 (0.0) | 1.0 (0.0) | 0.9772<br>(0.0168) | 0.9955<br>(0.0046) | 0.9913<br>(0.0098) | 0.9646<br>(0.0151) |
|  | Generalize<br>d | 0.9904<br>(0.0144) | 0.9646<br>(0.0381) | 0.9128<br>(0.0774) | 0.9915<br>(0.0156) | 0.9323<br>(0.1255) | 0.926<br>(0.936) |
|  | Focal | 0.9934<br>(0.0099) | 0.9706<br>(0.0385) | 0.9313<br>(0.0731) | 0.97<br>(0.0565) | 0.9634<br>(0.0513) | 0.936<br>(0.0648) |
|  | Weighted | 0.9961<br>(0.0035) | 0.9912<br>(0.007) | 0.9689<br>(0.0177) | 0.98<br>(0.0148) | 0.9628<br>(0.0198) | 0.9552<br>(0.0139) |
| Mob<br>ilene<br>tV3 | CAE | 0.9919<br>(0.0094) | 0.9919<br>(0.0078) | 0.9654<br>(0.0226) | 0.9656<br>(0.0351) | 0.9505<br>(0.0468) | 0.8576<br>(0.0517) |
|  | IESS | 0.9999<br>(0.0002) | 0.9998<br>(0.0003) | 0.9799<br>(0.0187) | 0.9957<br>(0.0065) | 0.9886<br>(0.0194) | 0.9942<br>(0.0071) |
|  | Generalize<br>d | 0.9921<br>(0.0095) | 0.9403<br>(0.0747) | 0.8307<br>(0.1389) | 0.9912<br>(0.0111) | 0.9204<br>(0.1109) | 0.8006<br>(0.1248) |
|  | Focal | 0.9989<br>(0.0015) | 0.9901<br>(0.0129) | 0.8416<br>(0.0636) | 0.9971<br>(0.0025) | 0.9748<br>(0.018) | 0.6833<br>(0.127) |
|  | Weighted | 0.9957<br>(0.0039) | 0.9894<br>(0.0081) | 0.9468<br>(0.0263) | 0.9829<br>(0.016) | 0.9645<br>(0.0297) | 0.8918<br>(0.0414) |
| Den<br>seNe<br>t | CAE | 0.929<br>(0.0563) | 0.9433<br>(0.0423) | 0.9456<br>(0.0333) | 0.9846<br>(0.0082) | 0.978<br>(0.0126) | 0.9445<br>(0.041) |
|  | IESS | 0.9969<br>(0.0047) | 0.9939<br>(0.0098) | 0.985<br>(0.0235) | 0.9964<br>(0.0019) | 0.9943<br>(0.0033) | 0.9337<br>(0.0393) |
|  | Generalize<br>d | 0.9278<br>(0.0495) | 0.6919<br>(0.0677) | 0.852<br>(0.0482) | 0.8012<br>(0.0793) | 0.5004<br>(0.1595) | 0.6506<br>(0.123) |
|  | Focal | 0.9886<br>(0.0128) | 0.9165<br>(0.0938) | 0.766<br>(0.114) | 0.9993<br>(0.0015) | 0.9937<br>(0.0127) | 0.898<br>(0.0749) |
|  | Weighted | 0.961<br>(0.023) | 0.9347<br>(0.0251) | 0.9367<br>(0.0215) | 0.9711<br>(0.0096) | 0.9352<br>(0.0177) | 0.9052<br>(0.0352) |

Table 3 illustrates that the usage of frequency bands can either be advantageous or disadvantageous for multi-class seizure detection depending on the model. For high-performing models, such as ResNet50 and DenseNet, no significant difference was observed in the weighted average performance when using raw signal inputs or frequency bands. However, models such as CNN2D+LSTM and MobileNetV3 showed significantly higher performance when using raw signal inputs as opposed to frequency bands.

Our results further demonstrate that performance in classifying specific seizure types depends on the selection of an appropriate feature extractor and model. The CAE enhanced the performance when raw signal inputs were used, specifically with the CNN2D+LSTM, ResNet50, and MobileNetV3 models. Contrarily, DenseNet exhibited improved performance when the frequency bands were used as feature extractors for EEG signals of CAE. Interestingly, IESS showed no significant changes in the AUC, regardless of the feature extractors and models used. GN showed similar performance irrespective of whether a feature extractor was used for models such as ResNet50 and MobileNetV3. However, GN displayed higher performance when raw signal was used with CNN2D+LSTM and DenseNet. Finally, FC showed comparable and non-significant performance, irrespective of the feature extractors and models used.

## DISCUSSION

This study marks a significant step forward in the application of real-time seizure detection in pediatric epilepsy cases, an area for which such techniques have not yet been applied. Although numerous attempts have been made to detect seizures, our study underscores the need to manage this issue in real-time, a feat that was previously regarded as challenging. Furthermore, the unique neurophysiological patterns in children and adolescents present a particular challenge, and the ability of our model to handle these intricacies demonstrates its potential and versatility.

Many seizure detection studies rely on pre-cut seizure signals.^16–18^ Tang et al.^24^ used the sliding window method; however, the non-overlapping window shifts compromised detection resolution, impeding timely intervention. Shawki et al.^12^ applied overlapping sliding windows in their approach, but the method demonstrated a comparatively lower seizure detection performance.

We achieved superior performance with one of our models, ResNet-short LSTM with 12 s window size. This model displayed an AUROC of approximately 0.98 in detecting any seizures across four baselines (Table 2). This highlights the applicability of our real-time seizure-detection techniques to real-world pediatric datasets, even in complex and diverse clinical settings. Detecting the seizure classes IESS and GN posed a significant challenge, as was reflected by their comparatively lower performance compared to other seizure types. The brief sequences of IESS events complicate their detection, and the heterogeneity of GN, which encompasses diverse features such as head drop and atonic seizures, adds to this complexity. Our results indicate that adopting shorter sliding windows is advantageous for detecting IESS due to its fleeting nature. In the context of FC, the window size did not significantly influence the detection performance. Despite the inherent challenges of seizure-specific detection, our model demonstrated robustness across various configurations and window lengths, underscoring its versatility in detecting pediatric seizure types.

Furthermore, our model demonstrated an impressive ability to accurately classify multiple types of seizures, demonstrating an overall performance score of more than 0.95 in classifying any kind of seizure (Table 3). This represents a critical advantage as it allows clinicians to rapidly and accurately diagnose and differentiate various seizure types, ultimately facilitating the provision of more personalized care.

A notable result of our study was the heterogeneous performance in classifying certain seizure types, which depended on the model and feature extractor. The findings in Table 3 suggest the need for careful selection of feature extractors based on the specific seizure type and chosen machine learning model. A remarkable finding was the insignificant changes in IESS detection performance, regardless of the model and feature extraction, which indicated that the classification task was relatively straightforward despite the challenges of detecting IESS seizure events. Furthermore, the multi-classification model performed flawlessly (AUC of 1.0) on IESS. The unique challenge with IESS is its detection rather than classification, as IESS has a distinct, brief characteristic that can make initial detection challenging. However, once detected, the distinctive nature of these spasms makes their classification relatively straightforward, which explains the exceptional performance of the model. Furthermore, the GN class showed relatively low performance due to the heterogeneous nature of the seizure class discussed above.

Although our findings are promising, we acknowledge the limitations inherent to the study’s retrospective design and the challenges of validation in a real-world setting. Although we performed two-stage real-time seizure detection and classification, these two processes were not implemented end-to-end fashion in the current model. Therefore, there is room for improvement. Future developments could aim to incorporate simultaneous and end-to-end real-time detection and classification [Figure 1 (b)], which would further enhance the clinical applicability and efficiency of our model. Further prospective studies are necessary to confirm our results and establish the real-time applicability of the model in other clinical environments. Nevertheless, our study established a foundation for developing a system for real-time seizure detection and classification, particularly for pediatric population.

Our study underscores the vast potential of deep learning and artificial intelligence in transforming the pediatric neurology landscape, particularly in terms of early detection and classification of seizures. With continued refinement, these systems promise a future in which epilepsy management can be greatly enhanced, providing patients with rapid, effective, and personalized care.

In conclusion, we validated that our real-time seizure detection framework and model could be applied to detect seizures in a real-world setting using EEGs from a pediatric neurology ward. The real-time seizure detection results with models benchmarked from previous studies showed that this seizure detection framework exhibit good performance for screening out seizures and thus could be applied in a realistic setting. Furthermore, multiple models tested in this work can be used to classify multiple seizure types, with good performance for different pediatric seizure types.

## DATA AVAILABILITY

Data collected for this study cannot be made publicly available to others due to the regulations for access to Severance Children’s Hospital’s medical data. The regulation is based on the Institutional Review Board of Severance Hospital (IRB 4-2023-0967). Analysis code for the seizure detection at the following repository: https://github.com/AITRICS/EEG_real_time_seizure_detection

## DECLARATION OF INTERESTS

The authors declare no conflict of interest.

## ACKNOWLEDGEMENTS

This study was supported by Basic Science Research Program through the National Research Foundation of Korea (NRF) funded by the Ministry of Education (NRF-2022R1A2C1012522), a grant of the Korea Health Technology R&D Project through the Korea Health Industry Development Institute (KHIDI) funded by the Ministry of Health and Welfare, Republic of Korea (grant number: HI21C1659), and the Team Science Award of Yonsei University College of Medicine (6-2021-0007).

---

**Supplemental Figure 1.**
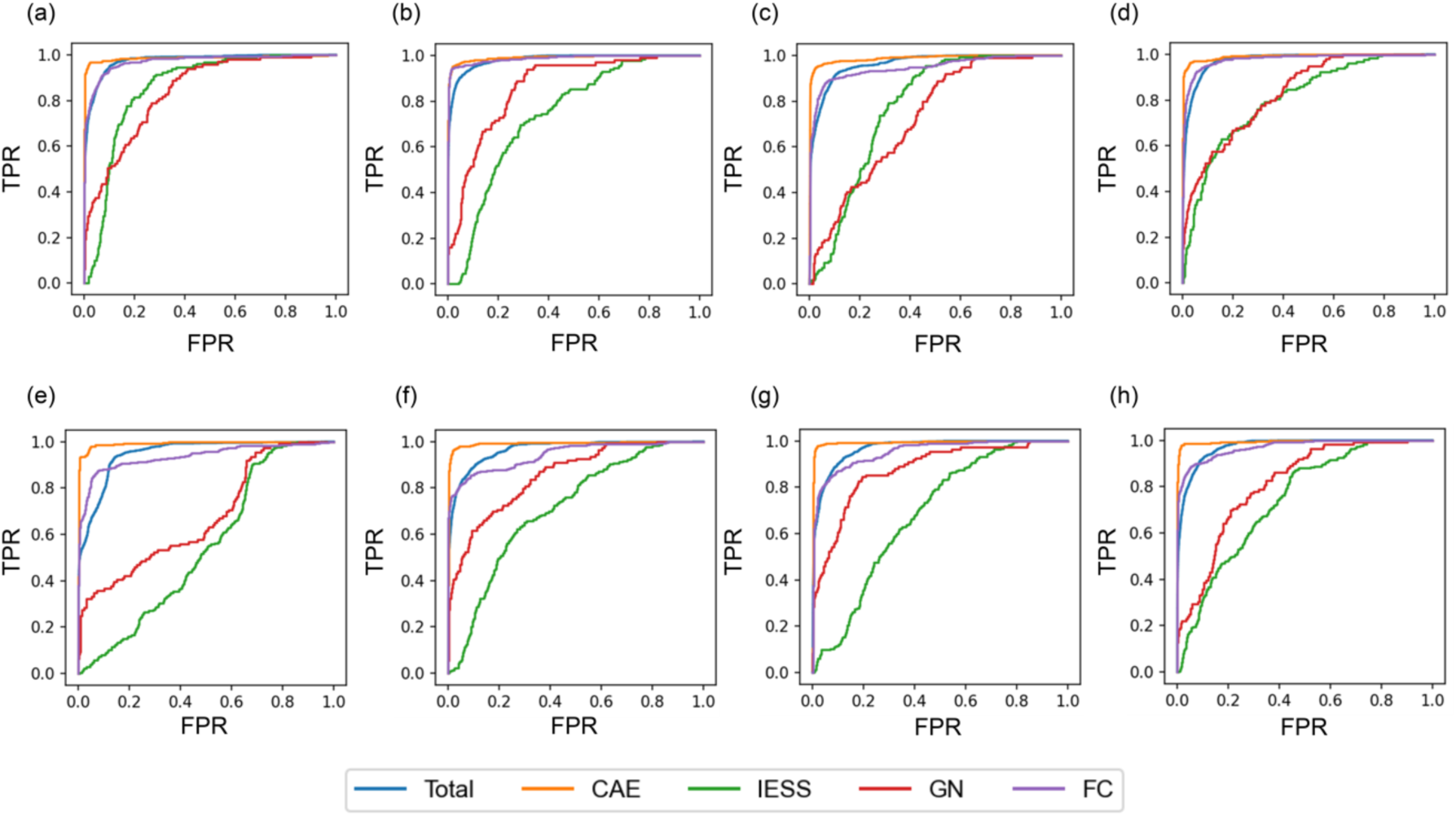
Area Under the Curve (AUROC) plots for binary seizure detection by different models and sliding window sizes (4 s figures (a)-(d), 12 s (e)-(h)): (a), (e) CNN2D+LSTM, (b), (f) ResNetshort+LSTM, (c), (g) ResNetshort+Dilation+LSTM, (d), (h) MobileNetV3short+LSTM.

**Supplemental Figure 2.**
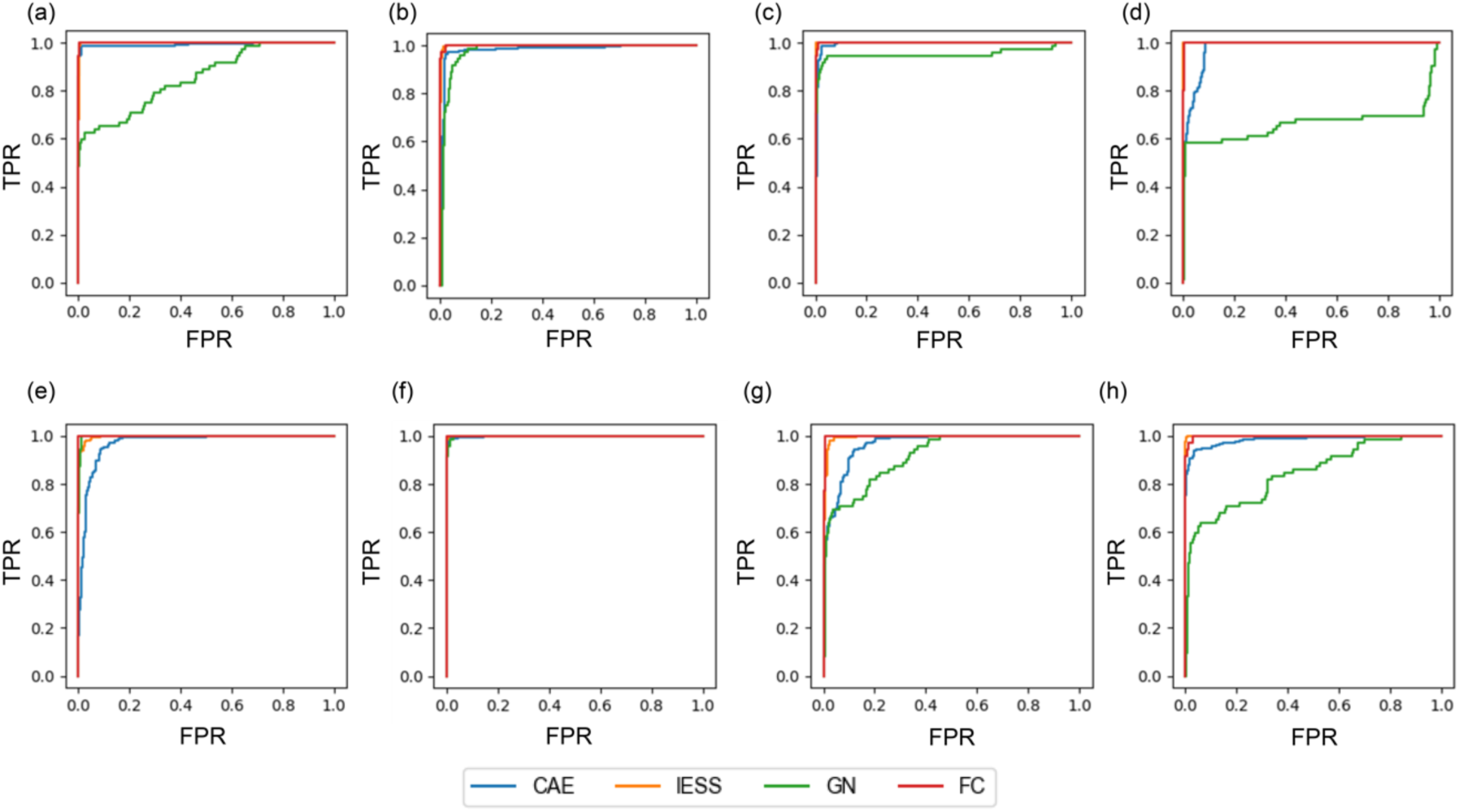
Area Under the Curve (AUROC) plots for multi-class seizure classification by different models and by feature extractor (raw signal (a)-(d), power spectral density (frequency bands) (e)-(h)): (a), (e) CNN2D+LSTM, (b), (f) ResNet50, (c), (g) MobileNetV3, (d), (h) DenseNet.

## Notes

**Financial support:** This study was supported by Basic Science Research Program through the National Research Foundation of Korea (NRF) funded by the Ministry of Education (NRF-2022R1A2C1012522), a grant of the Korea Health Technology R&D Project through the Korea Health Industry Development Institute (KHIDI) funded by the Ministry of Health and Welfare, Republic of Korea (grant number: HI21C1659), and the Team Science Award of Yonsei University College of Medicine (6-2021-0007).

### Competing Interest Statement

The authors have declared no competing interest.

### Author Declarations

The Institutional Review Board of Severance Hospital gave ethical approval for this work (IRB 4-2023-0967).

